# Development and Clinical Validation of Swaasa AI Platform for screening and prioritization of Pulmonary TB

**DOI:** 10.1101/2022.09.19.22280114

**Authors:** Gayatri Devi Yellapu, Gowrisree Rudraraju, Narayana Rao Sripada, Baswaraj Mamidgi, Charan Jalukuru, Priyanka Firmal, Venkat Yechuri, Sowmya Varanasi, Venkata Sudhakar Peddireddi, Devi Madhavi Bhimarasetty, Sidharth Kanisetti, Niranjan Joshi, Prasant Mohapatra, kiran Pamarthi

**Affiliations:** Andhra Medical College, Visakhapatnam, India; Salcit Technologies, Jayabheri Silicon Towers, Hyderabad India; C-CAMP; Department of Computer Science, University of California, Davis

**Keywords:** Pulmonary Tuberculosis (PTB), Cough signature, Convolutional Neural Network (CNN), Feedforward Artificial Neural Network (FFANN), Machine learning

## Abstract

Acoustic signal analysis has been employed in various medical devices. However, studies involving cough sound analysis to screen the potential Pulmonary Tuberculosis (PTB) suspects are very few. The main objective of this cross-sectional validation study was to develop and validate the Swaasa AI platform to screen and prioritize at risk patients for PTB based on the signature cough sound as well as symptomatic information provided by the subjects. The voluntary cough sound data was collected at Andhra Medical College-India. An Algorithm based on multimodal Convolutional Neural Network (CNN) architecture and Feedforward Artificial Neural Network (FFANN) (tabular features) was built and validated on a total of 567 subjects, comprising 278 positive and 289 negative PTB cases. The output from these two models was combined to detect the likely presence (positive cases) of PTB. In the clinical validation phase, the AI-model was found to be 86.82% accurate in detecting the likely presence of PTB with 90.36% sensitivity and 84.67% specificity. The pilot testing of model was conducted at a peripheral health care centre, RHC Simhachalam-India on 65 presumptive PTB cases. Out of which, 15 subjects truly turned out to be PTB positive with a Positive Predictive Value of 75%. The validation results obtained from the model are quite encouraging. This platform has the potential to fulfil the unmet need of a cost-effective PTB screening method. It works remotely, presents instantaneous results, and does not require a highly trained operator. Therefore, it could be implemented in various inaccessible, resource-poor parts of the world.

## Introduction

Tuberculosis (TB) is the world’s second leading airborne infectious disease after COVID-19. Unlike COVID the causative agent of TB is a bacterium, *Mycobacterium tuberculosis* (MTB). However, both the infections mainly affect the respiratory system. Although the bacteria have the capability to cause infection in various body parts. Pulmonary Tuberculosis (PTB) along with Extra pulmonary tuberculosis (EPTB) are the active form of infection, which displays symptoms such as fever, night sweat, weight loss and cough ^1,2^. In 2020, the World Health Organization globally reported nearly 10 million active TB cases and 1.5 million TB related mortalities. Although, TB is curable and preventable, the number of active cases is still high in various low income, developing countries including India. An active PTB patient can infect nearly ten to fifteen people every year ^3^. Currently there are several methods for diagnosing presumptive as well as active PTB cases, such as sputum staining, chest X-ray (CXR) and sputum cartridge based nucleic acid amplification test (CB-NAAT) or sputum GeneXpert test. However, all these methods are very expensive, require proper lab setting and trained technicians. Therefore, quick, and inexpensive mass screening methods ^4^ are required for reducing the transmission of infection by providing timely diagnosis followed by appropriate treatment regime ^5,6^.

Cough is a common symptom of respiratory disease and is caused by an explosive expulsion of air to clear the airways ^7^. It is a significant feature of pulmonary tuberculosis and results in the release of airborne particles into the environment ^8,9^. There are commonly two types of coughs i.e., wet and dry. Coughs are classified as wet when they have auditory characteristics that are suggestive of mucus, and dry when there is no discernible wetness ^10^. It has also been postulated that the glottis behaves differently under different pathological conditions, which makes it possible to distinguish coughs originating from different underlying conditions such as asthma, bronchitis, and pertussis (whooping cough) ^11^. Since coughing is a dominant symptom of PTB, there are reports which suggest that the coughing sound of an individual with pulmonary TB has some unique characteristic features that distinguish the diseased condition from the normal scenario ^12–14^. Still, a lot of research is needed to fully explore and decode the information contained in the cough sound to use it as an indicator of the underlying disease.

The recent application of Artificial Intelligence (AI) and advances of ubiquitous computing for respiratory disease prediction has created an auspicious trend and myriad of future possibilities in the medical domain ^15–17^. There is an expeditiously emerging trend of Machine learning (ML) and Deep Learning (DL)-based algorithms exploiting cough signatures ^18^. Cough analysis approaches are primarily subjective and are affected by the limitations of human perception. Audiometric analysis of cough (digital signal) provides essential information about characteristics of cough sounds in different respiratory pathological conditions. Several studies have been conducted in the past to collect and analyse cough sound data for PTB pre-screening and triaging using mobile devices. However, there are some missing links in terms of selecting the subjects, collecting the cough data and lack of proper technical/ clinical validations to scale up these tools for mass screening of PTB subjects ^8,9,13,19,20^.

Our study provides a holistic approach by developing, validating, and testing the “Swaasa AI platform” to screen and prioritize the potential PTB cases. It is a SaMD (Software as a Medical Device) that evaluates respiratory health using a 10-second cough sound recording, serving as a quick Point of Care tool. It effectively prioritizes at-risk patients for molecular testing when used as a screening and triaging tool. As opposed to majority of the previous reports that utilized the crowdsource cough sound database for training their model, we have conducted the data collection from 567 unique subjects for our model derivation as well as validation phase in a proper clinical setting. Hence, our data have cough recordings collected from various unique subjects to build a robust model. Unlike others, we have trained two parallel models i.e., Convolutional Neural Network (CNN) model with Mel spectrograms and Feedforward Artificial Neural Network (FFANN or tabular) model with primary as well as secondary features and merged the final layer to build a combined logic. In the validation phase, the AI-model was found to be 86.82% accurate in detecting the likely presence of PTB with 90.36% sensitivity and 84.67% specificity. Therefore, it satisfies the specificity (70%) and sensitivity (90%) criteria set by the World Health Organisation (WHO) for a community-based mass TB screening test ^14^. The results obtained by the model are very promising with a scope to make it scalable for quick, cost-effective, and non-invasive screening of PTB cases. A large-scale study will further help us to improvise the accuracy of the platform for making it more reliable for screening genetically diverse subjects under different environmental conditions.

## Materials and Methods

### Sample size estimation

To calculate the adequate sample size for our study, we used a simple formula that required us to select appropriate values for several assumptions. The formula used was n=Z^2^*P(1−P)/d^2^, where n represented the sample size, Z was the statistic corresponding to the level of confidence, P was the expected prevalence, and d was the precision corresponding to the effect size ^21^. By using this formula, we were able to determine an appropriate sample size for our study. These many number of subjects were appropriate for validating if the device could detect PTB respiratory conditions with a 90% sensitivity on considering a 1% error for a 95% confidence interval (CI) and a prevalence of 0.75% as the highest prevalence of PBT in India is 0.747% (747 per 100,000 population) ^22^. In total 567 subjects were recruited, out of which 50.9% subjects were classified as controls. The control group consisted of both healthy individuals and those who displayed respiratory disease symptoms but tested negative for PTB via CB-NAAT. These respiratory conditions included asthma, Chronic Obstructive Pulmonary Disease (COPD), Interstitial lung disease (ILD), and pneumonia. The number of TB records were calculated based on disease prevalence. In order to avoid potential bias in the model, it was trained using an equal number of TB and non-TB records.

### Data Collection

The cough data has been collected at Andhra Medical College (AMC), Visakhapatnam, India as part of the clinical study “Swaasa Artificial Intelligence Platform for detecting the likely presence of Pulmonary Tuberculosis”. The study was registered with Clinical Trials Registry-India (CTRI/2021/09/036609) on 17^th^ September 2021. The methods were performed in accordance with relevant guidelines and regulations and approved by AMC-Institutional Ethics Committee (IEC). Written informed consent was taken from all the enrolled subjects. After getting the informed consent, the patient’s demographic details and vitals were collected. The patients were also interviewed as per the Part I of the St. George’s Respiratory Questionnaire (SGRQ) ^23^, which primarily covers the symptoms they’ve had experienced within the past few months or year. This was followed by cough sound collection by trained health care personnel via a smartphone (Android or iPhone). To ensure the highest quality data for analysis, several factors were taken into consideration before recording. The person being recorded was given specific instructions to sit comfortably in a quiet place, hold the recording device (which included smartphones and tablets from various manufacturers) 4-8 inches away from their mouth, and maintain a 90-degree angle with their face. They were also instructed to take a deep breath and cough 2-3 times until the recording stopped, which lasted for 10 seconds. However, since the collected data was from varying environments and a variety of devices, it was important to control as many potential variables as possible. Therefore, noise filtering was applied using a noise reduction algorithm. This algorithm calculated the ratio of the power of observed signals at two microphones for smartphones with two or more built-in microphones, and then calculated the spectral gain function based on the power level ratio using the sigmoid function. The result was a denoised audio recording. For smartphones with a single built-in microphone, noise filtering was not applied during the recording. Instead, a noise removal technique was applied during pre-processing. This involved subtracting the noise audio clip (which contained background noise such as electronic noise, multiple people talking, and fan sound) from the signal audio clip (which contained the cough). The noise removal technique isolated the signal using Fast Fourier Transform, removing the background noise and resulting in a cleaner recording. Valid coughs were detected using a cough/non-cough classifier, which screened the dataset for coughs with high background noise. If a recording did not meet the minimum required valid coughs, a message would appear on the mobile screen instructing the person to give another recording following the instructions. Overall, these processes standardized the dataset, making it suitable for analysis.

During the audio recording process, we implemented several safety measures to prevent the transmission of disease. All subjects were required to wear a surgical mask while providing the audio recordings, in order to limit the spread of germs through water droplets during coughing. After each recording, the phone used for recording was cleaned using one of three methods. A disinfectant wipe was used to clean the phone, or alternatively, a damp microfiber cloth dipped in soapy water was used if the phone was waterproof. Another option was to use a mobile sanitizer to clean the phone. These measures helped to maintain a clean and safe environment during the data collection process.

Following the cough sample collection, patients were subjected for CB-NAAT and chest X-Ray (CXR P/A) view for diagnosis of PTB. The data distribution across different gender and age groups is presented in Figure 1. The inclusion criteria were that a patient must be of (a) age ≥ 18 years and should display (b) symptoms suggestive of PTB (presumptive PTB). Whereas patients with (a) age ≤ 18 years and who were (b) on ventilators support were completely excluded from the current study. COVID precautionary and infection control measures were followed strictly.

**Figure 1:**
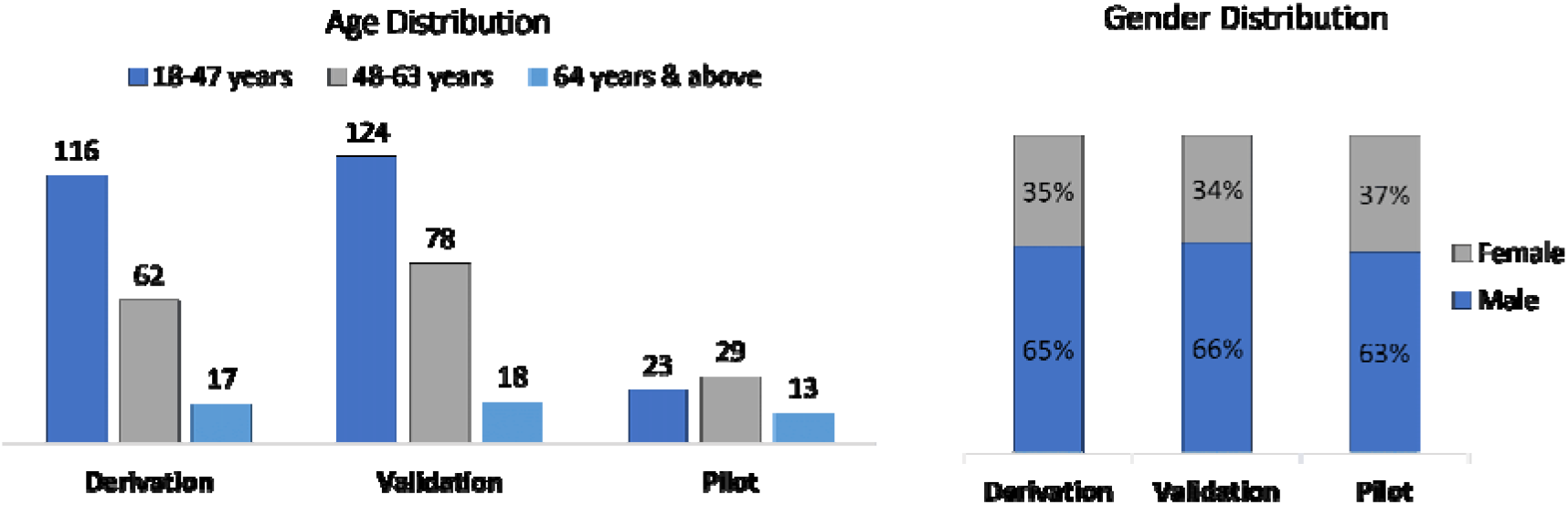
Data distribution in the derivation phase, validation phase and pilot testing.

### Model development and training

In the Phase 1 (Derivation phase) of the study, we aimed to develop and train a machine learning-based model for the detection of pulmonary tuberculosis (PTB) using cough sounds. The objective of this phase was to quantify the technical as well as analytical performance of the device by establishing a unique cough signature for PTB. A total of 195 PTB positive cases were recruited for the derivation phase, and audio cough recordings were collected. In addition, 152 non-PTB subjects were also included to train the model to distinguish between PTB condition and normal healthy subjects as well as other respiratory disease scenarios.

Event extraction was carried out from the collected audio cough records using the moving window signal standard deviation technique ^24^. A cough/non-cough classifier was used to segregate the events into actual coughs and non-coughs such as silence, speech, fan sounds, vehicle sounds like horn, and noise. A total of 3102 cough events were extracted at this step. The features were extracted from the time as well as frequency domain of each cough event. The important time domain features that were taken into consideration were Zero crossing rate (ZCR) and Energy. The frequency domain features which were utilized for data analysis are MFCC, Spectral centroid, Spectral bandwidth, and Spectral roll-off ^25^. The features were extracted for each frame within the cough signal. Each frame was typically about 20 ms in duration. The cough event duration can vary from anywhere between 200 ms to 700 ms.

The total features extracted were 209, that includes age, gender, 120 Mel Frequency Cepstral coefficients (40 MFCC, 40 first derivatives of MFCC, 40 second order derivatives of MFCC), 9 spectral features (spectral centroid, spectral roll-off, spectral bandwidth, dominant frequency, spectral skewness, spectral kurtosis, spectral crest, spectral spread and spectral entropy), 33 chroma features (11 chroma, 11 first derivatives of chroma, 11 second derivatives of chroma), 18 contrast features (6 contrast, 6 first derivatives of contrast, 6 second derivatives of contrast), 15 tonnentz features (5 tonnentz, 5 first derivatives of tonnentz, 5 second derivatives of tonnentz), 3 Zero-crossing rate (ZCR, first derivatives of ZCR, second derivatives of ZCR), 3 Energy (Energy, first derivatives of energy, second derivatives of energy), 3 skewness (skewness, first derivatives of skewness, second derivatives of skewness), 3 kurtosis (kurtosis, first derivatives of kurtosis, second derivatives of kurtosis). On these features, we did correlation analysis and recursive feature elimination (RFE) to rank the feature according to their importance. Correlation-based feature selection was used to reduce the feature size from 209 to 170, and highly correlated features were removed to prevent overfitting and improve the performance of the model. Primary features include all the 170 features. The secondary features included age (categorized), gender, symptoms, cough type (dry/wet), and cough duration. The cough type is derived from the primary features and cough duration is derived from audio signal. The CNN model is trained with the Mel spectrograms of cough sounds. Whereas, both secondary and primary features were used to train the FFANN model.

The CNN model used in the study was based on transfer learning using Resnet-34 with imagenet for training on spectrograms. Whereas, the FFANN was utilized to process the tabular data. The FFANN consisted of two hidden layers, with 400 and 300 neurons, respectively. Each layer was followed by batch normalization. The selection of the number of layers, number of neurons in each layer, and the activation functions were determined using the Bayesian optimization method. The last fully connected layers of both models were removed, and a new fully connected layer (merged layer) was added to predict the final output. The merged layer consists of activation layers (Figure 2). This merging approach of the last layers of the two models was named the combined logic. When the model is uncertain about the likely detection of PTB as yes/no, it provides the output as inconclusive as displayed in the block diagram in Figure 3, wherein PTB likely indicates TB positive and PTB unlikely indicates TB negative condition.

**Figure 2:**
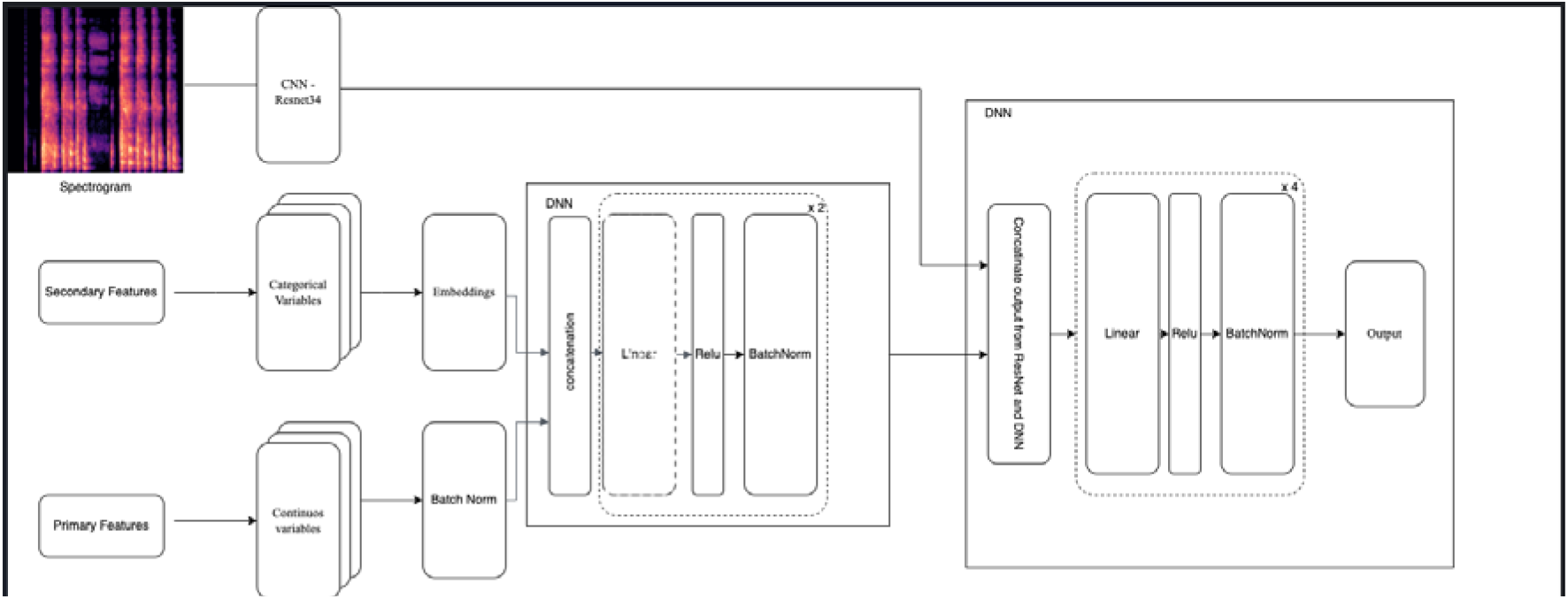
Illustration of the combined logic - combining Feedforward Artificial Neural Network (FFANN) model and Convolutional Neural Network (CNN) outputs

**Figure 3:**
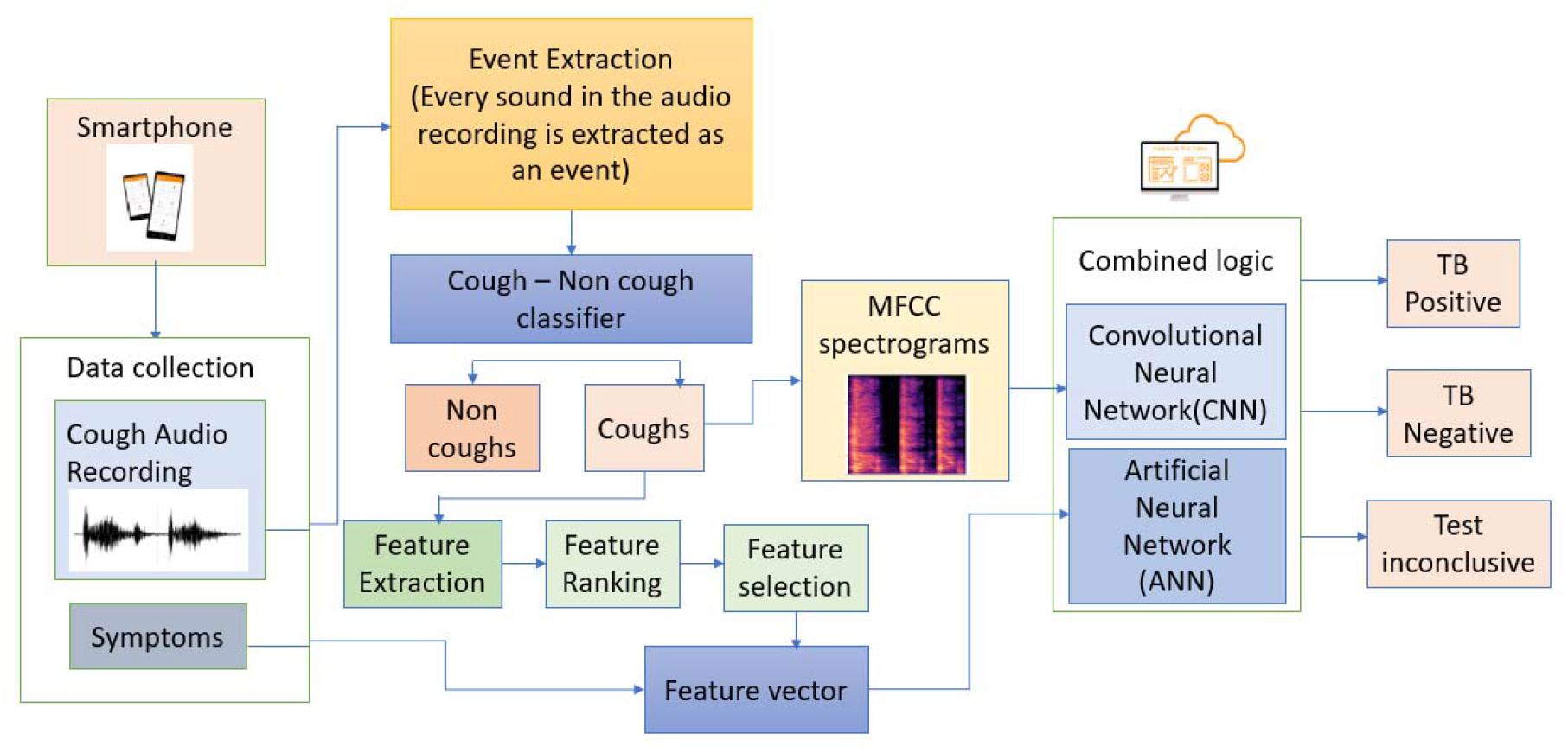
Block Diagram illustrating the flow of the TB prediction model

Overall, the primary and secondary features were used to train feedforward neural network models (tabular) and Mel spectrograms were used to train CNN and the combined logic approach was used to merge the outputs of the two models. The correlation-based feature selection was also used to improve the performance of the model.

During our study, we split the initial dataset into 80% training and 20% testing sets to assess the performance of the machine learning model. In addition, we used the k-fold (K=10) cross-validation approach to divide the training dataset into k subsets of data, which allowed us to obtain a more robust estimate of the model’s performance.

### Clinical validation of the Model

After training the model, it was tested in the Phase 2 i.e., Clinical validation phase. A total of 220 presumptive PTB cases were recruited and subjected to the screening test using the model. The results were compared with diagnosis based on sputum CB-NAAT test or radiological diagnosis, which are considered classical gold standard diagnostic methods. A total of 98% of the subjects underwent sputum CB-NAAT testing for the diagnosis of TB. In cases where the results of CB-NAAT testing were unclear for the remaining 2% of subjects, a repeat test was performed. If the results remained inconclusive after the second test, fresh sputum samples were collected. Additional tests such as acid-fast bacilli (AFB) staining, and chest X-ray (CXR) were performed to confirm the TB status of the patients.

The consolidated test summary sheet was generated, which contained the results obtained from the classical gold standard diagnosis methods along with the model’s output. Both the results were then compared by a statistician.

### External validation of the Model

In Phase 3 (Pilot Phase), the model was externally validated to evaluate its effectiveness as a screening tool for PTB detection prior to clinical diagnosis. The sample size for this phase consisted of 65 individuals who were identified as presumptive PTB cases and recruited from a peripheral healthcare center, RHC Simhachalam. The effectiveness of the model was measured by calculating the ratio of patients truly diagnosed as PTB positive via standard lab-based diagnostic techniques to all those who were predicted to be PTB positive via the AI-based model. The diagnostic performance of the model was evaluated using metrics such as sensitivity, specificity, positive predictive value (PPV), negative predictive value (NPV), and accuracy. To assess the effectiveness of the model, a data analysis strategy similar to that used in phase 2 was employed. The AI-based model was compared with classical gold standard diagnostic methods, such as sputum CB-NAAT testing or radiological diagnosis, and the results were analysed using statistical methods.

### LIME Representation

In Local interpretable model-agnostic explanations (LIME) representation ^26^, the green part shows where the model reacted positively for a particular class and red parts highlights where it reacted negatively. It explains the prediction by presenting textual or visual artefacts that provide qualitative understanding of the relationship between the instance’s components (e.g., words in text, patches in an image) and the model’s prediction.

### Statistical analysis

The comprehensive evaluation of the model performance on the test set includes accuracy sensitivity, specificity, positive prediction value (PPV), negative predictive values (NPV) and ROC. To measure the variability around these parameters, we used 95% confidence intervals using the Clopper–Pearson method ^27^. To better understand the performance of the model in screening PTB subjects, we also calculated confusion metrics on the entire test set.

## Results

### Patient population in Model derivation phase

Cough sound data was collected from 195 subjects PTB positive subjects and 152 PTB negative subjects in the derivation phase. Among 195 subjects, 65% were male and 35% were female, with age ranging from 18 years to 64 & above. Subjects were confirmed with TB by standard diagnosis methods. In this phase multiple data points were collected from the subjects. Each data point was called a record. A total of 597 cough records were collected from 195 patients. The data was annotated with disease condition as PTB i.e., PTB likely as “yes”. For PTB unlikely, data representing other respiratory disease conditions was added from the pre-existing labelled datasets (collected as a part of earlier studies) in appropriate propositions ^25^.

The features listed in Table 1 depicts the mean value of the features extracted from individual frames, where we have considered normal as well as respiratory diseases data other than PTB from our previous validation study conducted at Apollo Hospitals, Hyderabad ^25^.

**Table 1:**
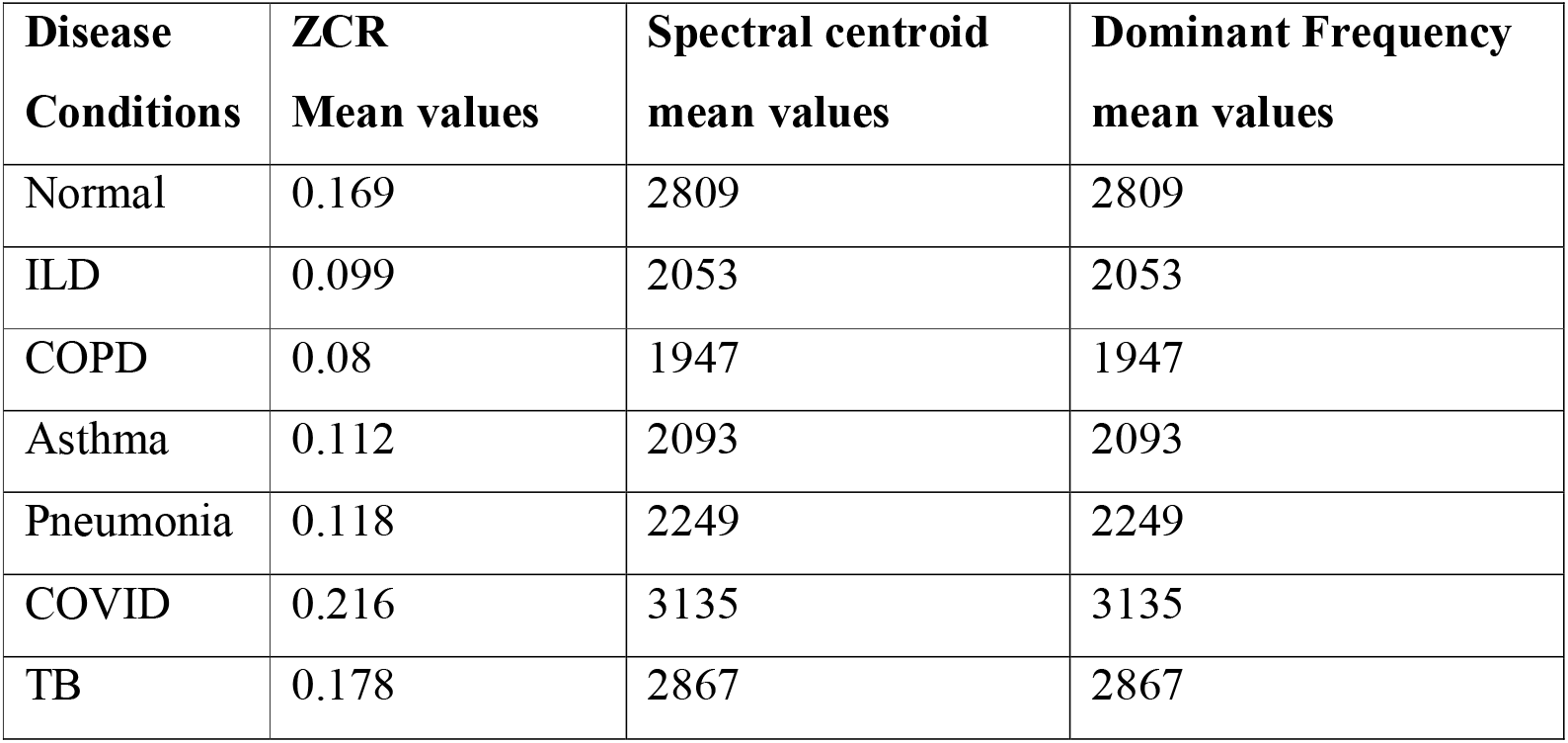
Table showing mean values of the Zero crossing rate (ZCR), spectral centroid and dominant frequency of various disease conditions.

### LIME data comparison

Spectral content is the distribution of audio signal based on its frequency w.r.t time, where high spectral content emphasizes that the energy of the cough bout remains same throughout the signal, whereas low spectral content corresponds to the conditions where the energy of the cough decreases with time. We observed that conditions like COPD and ILD carried very low spectral frequencies as compared to Asthma, which has a medium spectral frequency. On the other hand, we detected a very high spectral content for diseases where mucus accumulation in the airways and fluid accumulation in parenchyma region was present such as, PTB. Features like high spectral content brought uniqueness in the PTB cough, which differentiates it from the other respiratory diseases.

Thorough feature analysis of the cough sounds highlighted that the cough sounds could distinguish diseases. Variation in the cough duration and frequency distribution alters with the pathological conditions of the respiratory system ^9,28^.

We have enlisted a few examples of cough signatures, cough spectrograms and related LIME maps for different respiratory diseases, including PTB in Table 2. It is evident from the LIME maps that frequency distribution of the coughs is unique for each disease. To be specific, both Asthma & ILD have negative reactions in high frequency regions. TB has a positive reaction in the high frequency region and in the low frequency region. Normal cough signature is widely spread. However, it is not like other diseased conditions, where it has a strong patch around a given region. Similarly in the first column of the table, the variation of the amplitudes of the cough from bout to bout is different in coughs related to different diseases. As amplitudes vary, energy also varies from bout to bout.

**Table 2:**
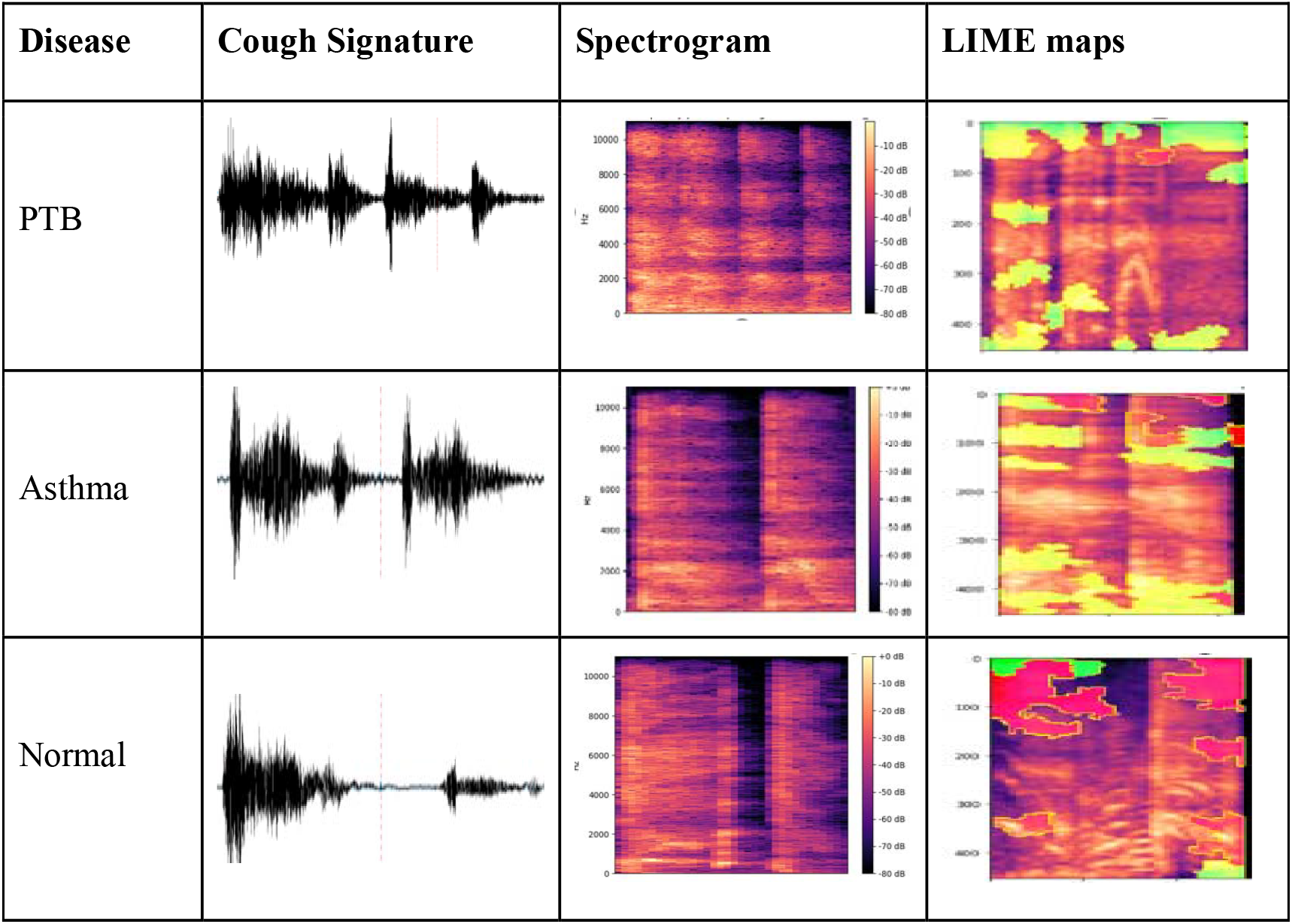
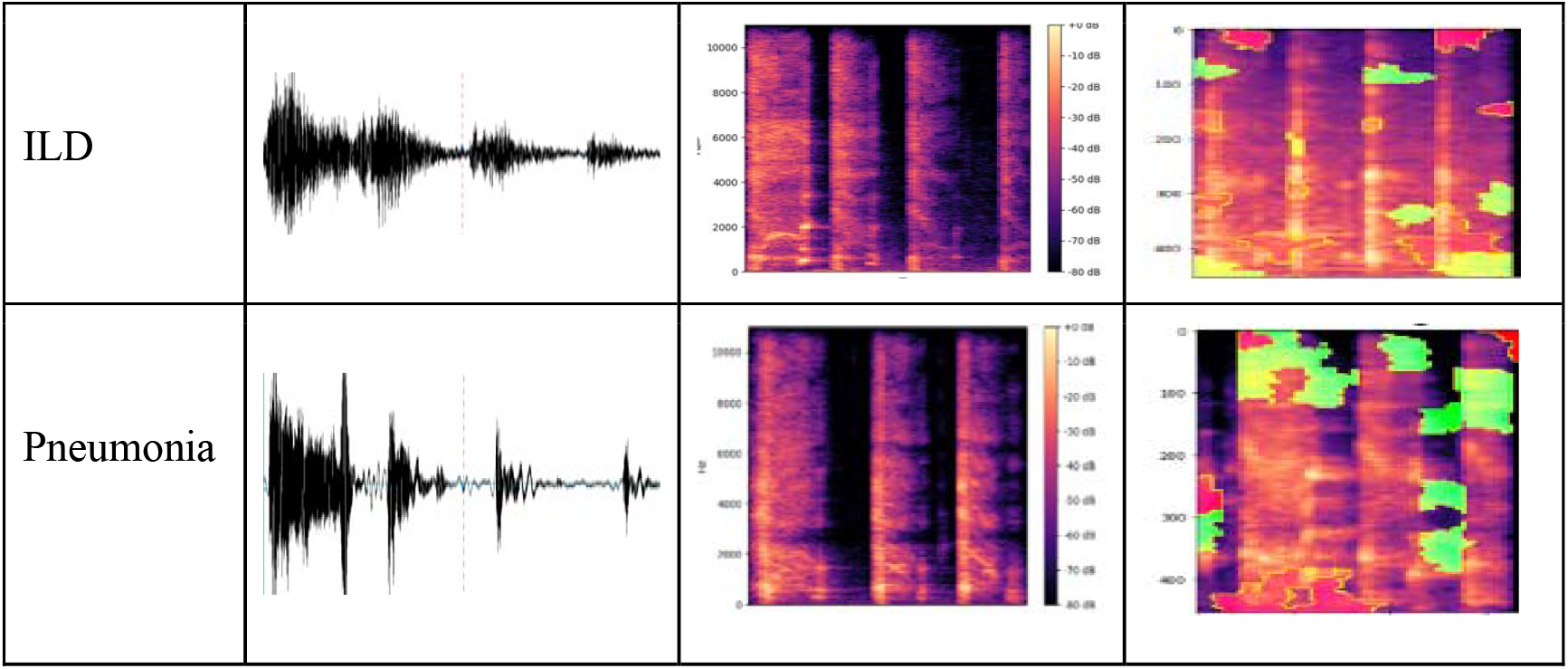
List of different respiratory diseases showing characteristic cough signature, cough spectrograms and related LIME maps.

From the feature analysis we conclude that PTB related cough has a unique signature, and it is captured by the features extracted from the cough, which can be identified by a machine learning model.

### Performance of Combined logic Model

Initially, the training data i.e., 3102 coughs which were extracted from 597 records collected from 195 subjects was internally divided into training and validation as required to build as well as optimize the model performance based on K-fold cross validation technique. The performance of our model was evaluated using k-fold cross validation, with k set to 10. The metric used for evaluation was the Area Under the Curve of the “Receiver Operating Characteristic (AUCROC) curve, which provides a measure of the model’s ability to distinguish between positive and negative samples. The obtained AUC score was 0.98, indicating that our model is highly effective in making accurate predictions. Figure 4 shows the representative ROC curve of the best performing fold among the 10 cross validation folds. In machine learning model, attribute like learning function, activation function were fixed for learning. Hence, the dataset was divided into subsets and the model was trained with each subset to validate the model.

**Figure 4:**
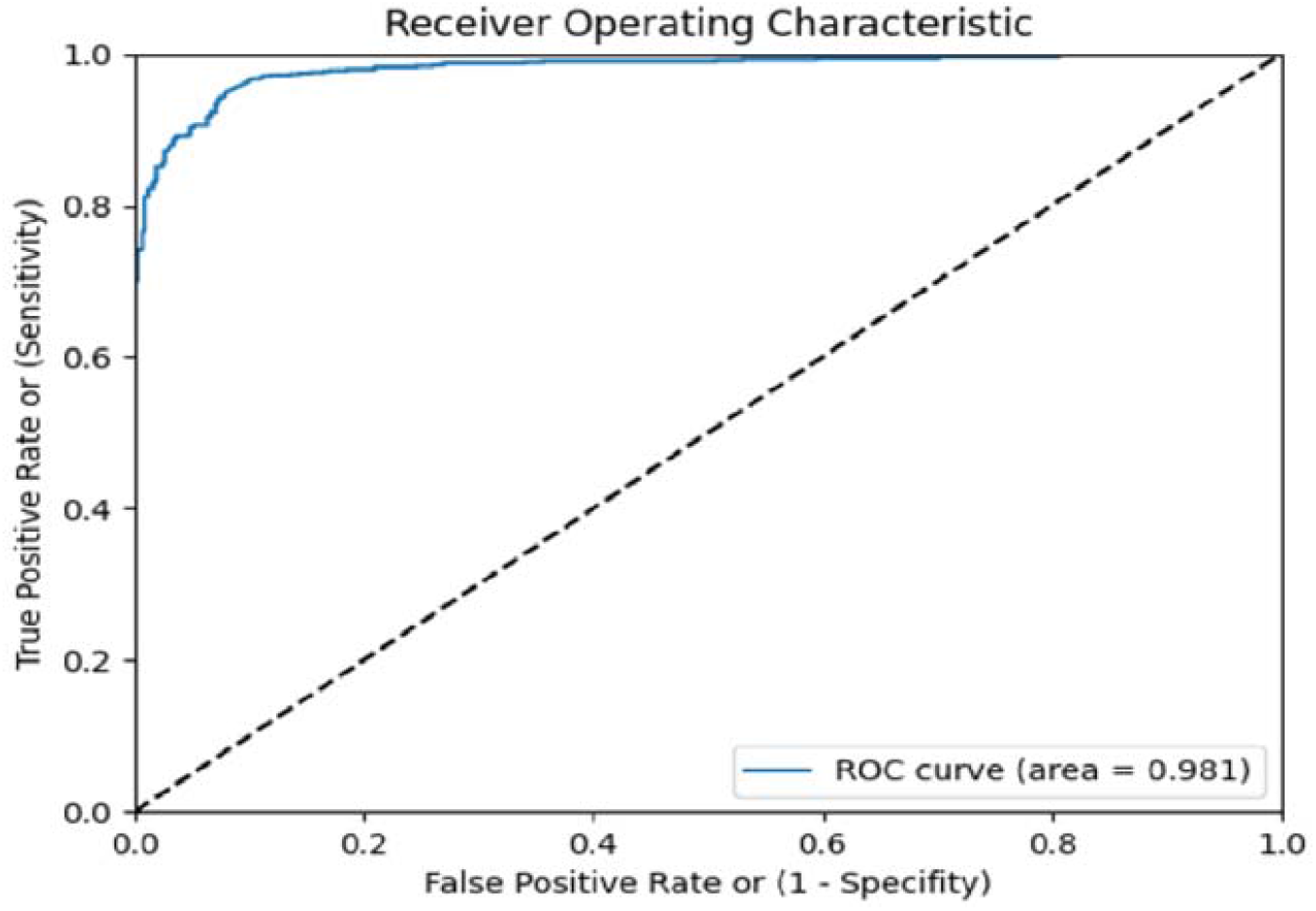
The representative graph for ROC curve, best among 10-fold validation of TB prediction model built using derivation data.

Further, the data collected in the derivation phase have been divided into 80% train and 20% test, when the test data was run through the classifier. We obtained four outcomes as enlisted in Table 3 i.e., 102 True positives (TP), 20 False Negatives (FN); 22 False Positives (FP) and 128 True Negatives (TN), that corresponds to 85% accuracy, 84% sensitivity and 85% specificity.

**Table 3:**
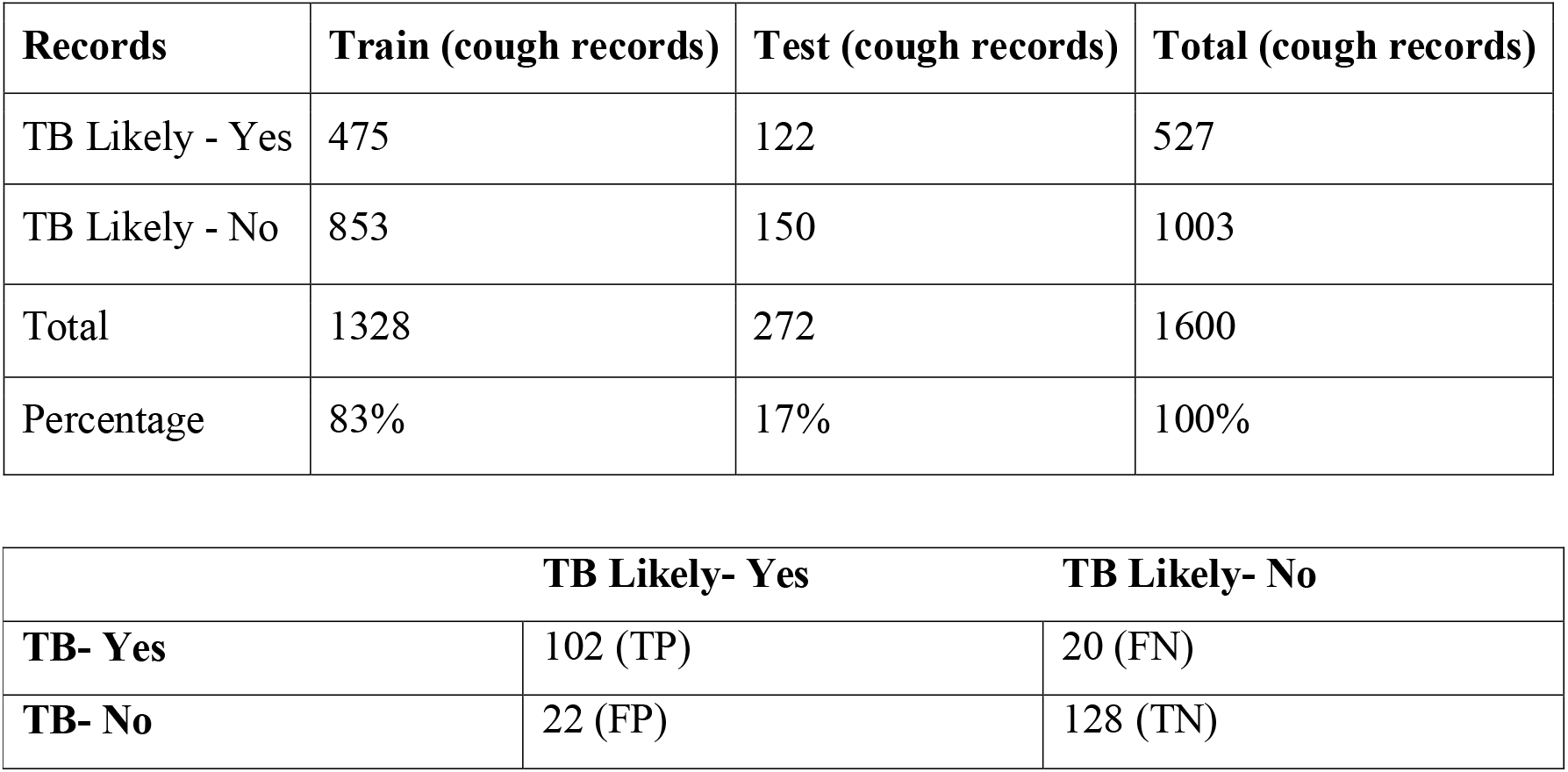
Data distribution into train data and test data along with final confusion matrix for the test data in derivation phase

A total of 220 subjects participated in the validation phase, out of which 83 subjects were found to be PTB positive and 137 subjects PTB negative by standard diagnostic methods. Only one cough record was collected from each subject in this phase. We achieved an AUC (Area Under the ROC Curve) of 0.94 (Figure 5). Confusion matrix for validation phase of the model is illustrated in Table 4, where the row represents the actual label, and the column represents predicted label. For the Validation phase we achieved an accuracy of 86.82% with 90.36% sensitivity and 84.67% specificity (Table 5).

**Table 4:** Confusion matrix for the validation phase

|  | TB Likely- Yes | TB Likely- No |
| --- | --- | --- |
| TB- Yes | 75 (TP) | 8 (FN) |
| TB- No | 21 (FP) | 116 (TN) |

**Table 5:** Performance metrics of the validation phase

| Statistic | Value | 95% CI |
| --- | --- | --- |
| Sensitivity | 90.36% | 81.89% to 95.75% |
| Specificity | 84.67% | 77.53% to 90.25% |
| Positive Likelihood Ratio | 5.90 | 3.95 to 8.79 |
| Negative Likelihood Ratio | 0.11 | 0.06 to 0.22 |
| Disease Prevalence | 37.73% | 31.30% to 44.49% |
| Positive Predicate Value | 78.12% | 70.54% to 84.19% |
| Negative Predicate Value | 93.55% | 88.20% to 96.57% |
| Accuracy | 86.82% | 81.62% to 90.99% |

**Figure 5:**
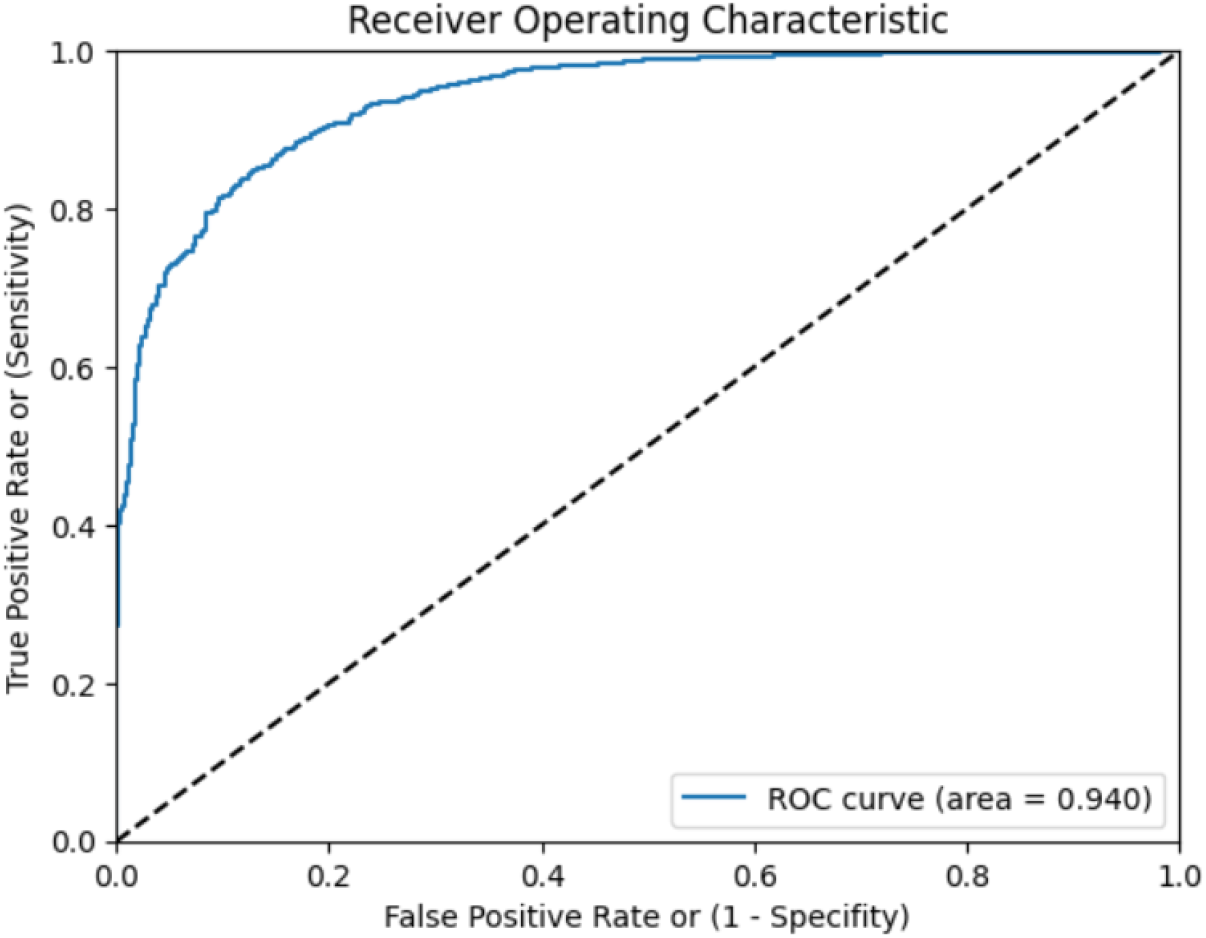
The provided graph shows the best ROC curve of a TB prediction model constructed using validation data.

### Model Output in the Pilot phase

Pilot testing was done on a total of 65 subjects. The patients approaching the testing centre with symptoms of cough suggestive of possible pulmonary tuberculosis are assessed for eligibility. Patient’s demographic details and vitals were collected and interviewed as per the SGRQ questionnaire. This is followed by cough sound collection by trained health care personnel.

Among 65 subjects, the model was able to identify 20 subjects as having a likely presence of TB. Out of these 20 subjects, 15 truly turned out to be TB positive with a Positive predictive value (PPV) of 75%. The confusion matrix for pilot testing phase is listed in Table 6. The model obtained a high AUC score of 0.90. Figure 6 shows the ROC curve of the model’s ability to distinguish between positive and negative samples.

**Table 6:** Confusion matrix for the pilot testing phase

|  | TB Likely- Yes | TB Likely- No |
| --- | --- | --- |
| TB- Yes | 15 (TP) | 5 (FN) |
| TB- No | 9 (FP) | 36 (TN) |

**Figure 6:**
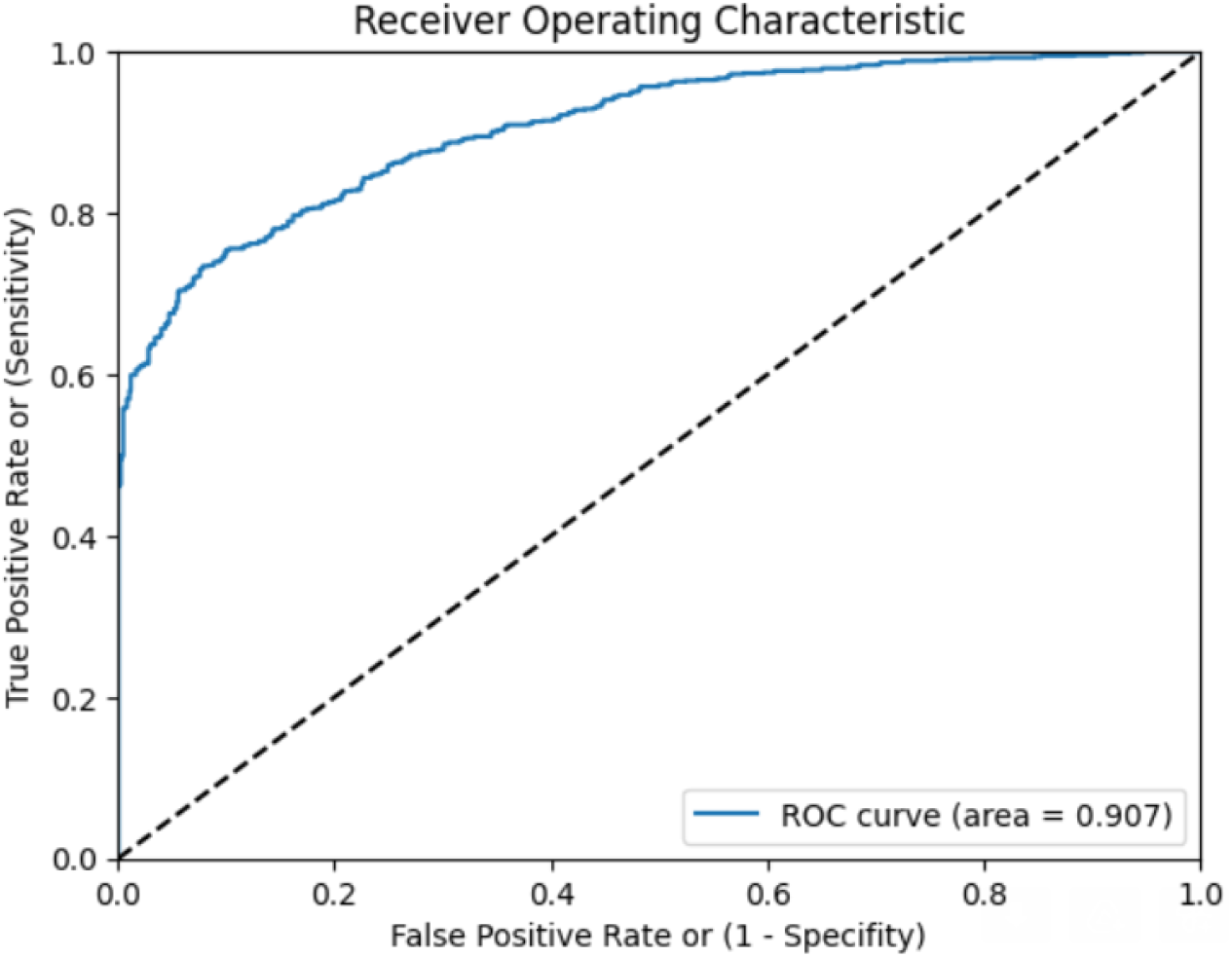
The provided ROC curve illustrates the performance of a TB prediction model constructed using pilot data

When compared to the existing classical methods, screening of PTB patients by the model saved a significant amount of time. Additionally, it does not require any trained professionals, the testing can be done by a community healthcare worker. The worker did not require any specific hardware or any other consumables. A smartphone with an internet connection is enough to conduct the test.

## Discussion

Several studies have been conducted in the past to deploy the information contained in the cough sound to detect and predict different disease outcomes such as Asthma, Pneumonia, COPD, bronchitis, and lung-cancer ^10,29–31^. Nowadays, due to the increasing COVID-19 cases, there has been a tremendous boost in the use of ML/DL frameworks to determine the presence of SARS-CoV-2 infection via cough sample analysis. This is because cough is one of the most prominent symptoms for the diseases that primarily affect the respiratory system. Numerous studies have shown that cough analysis can accurately predict COVID-19 ^32,33^. However, there are only a handful of clinical trials that emphasise the association of cough to the underlying Pulmonary TB condition ^12–14,20^. Most of the previously developed tools were utilizing the logistic regression methods to build the model. However, in the current study, we developed the model by combining the final output layers of the two separate models i.e., FFANN model (training input: primary and secondary features) and CNN model (training input: Mel spectrograms) because it gave us far better prediction outcome as compared to the either logical regression or CNN model used alone by other groups ^13,14,20^. We conducted the pilot screening on a comparatively large cohort, whereas previous studies were performed on a smaller scale. A pilot study conducted in Peru focused on analysing cough sounds for providing a foundation to support larger-scale studies of coughing rates over time for TB patients undergoing treatment ^20^. A similar cough sound analysis study was undertaken in South Africa for automatically classifying coughing sounds, which could be a viable low-cost and low-complexity screening method for PTB ^13^.

The approach of the current study is different with respect to the previously published data in terms of the amount of data collected to build and train the model. As compared to the maximum AUC of 0.94 achieved in a similar study upon utilizing only 23 features and with less dataset, we have utilized 170 features while training the model and achieved an AUC of 0.84 on a dataset comprising TB and non-PTB, where non-PTB includes other important diseases like Asthma, COPD, COVID-19, Pneumonia as well as healthy subjects ^14^. Having a greater number of latent features helps in distinguishing the signature better. Our model achieved an accuracy of 86.82% with 90.36% sensitivity and 84.67% specificity in the clinical validation phase. We conducted the pilot testing in a real primary care setting to test the accuracy of the tool. Upon deployment as a screening and triaging tool prior to molecular testing, the model was proven statistically effective in prioritizing at-risk patients for confirmatory testing. In the pilot phase also, the model achieved a positive prediction value of 75% in a clinical setup at a tertiary care hospital.

Considering the performance of the present diagnostic tests for PTB, our model’s technical and clinical validation results are quite encouraging, given the device is primarily intended to be used as a screening tool and helps in prioritizing and fast tracking the patients for subjecting them to the standard reference tests for confirmation of diagnosis of PTB.

During our study we observed that on an average 10 to 12 patients are diagnosed with extensive PTB with severe parenchymal damage, respiratory failure and poor lung function every month at a remote community health centre in India. Most of these patients belong to tribal areas. The delay in diagnosis is mainly due to lack of awareness, and social inhibitions in reaching a doctor or a peripheral health worker. We believe that this model will help in reducing the gap in accessibility for the much-needed population.

## Data Availability

All data produced in the present study are available upon reasonable request to the authors

## Data availability

Due to the nature of this research, participants of this study did not agree for their data to be shared publicly. However, the detailed analysis can be shared by NRS upon reasonable request.

## Author contributions

GDY and DMB defined study protocol, including the study design and methodology. NRS conceptualized the idea of using cough sounds for screening and diagnosing respiratory problems. GR performed literature review and data analysis. BM and CJ were involved in device development. VY created a value proposition for the device. VSP assisted in executing the project at AMC by providing all the resources and extending research capabilities. SV, SK and KP performed data analysis, sample size estimation and result analysis. PM provided subject matter expertise. GR and PF wrote the manuscript. All the authors provided intellectual inputs and helped in preparing the manuscript.

## Acknowledgement

This study is supported by the UK Government (British High Commission, New Delhi). This is a commissioned research report on commercial terms between C-CAMP and the UK Government (British High Commission, New Delhi). We would also like to acknowledge the team from Andhra Medical College Visakhapatnam, Government TB & Chest Hospital Visakhapatnam for all the support provided

## References

1. Al Lawati, R. et al. COVID-19 and Pulmonary Mycobacterium Tuberculosis Coinfection. Oman Med. J. 36, e298–e298 (2021).

2. Pai, M. et al. Tuberculosis. Nat. Rev. Dis. Prim. 2, 1–23 (2016).

3. WHO. Global tuberculosis report 2021. Geneva: World Health Organization; 2021. Licence: CC BY-NC-SA 3.0 IGO. (2021).

4. WHO. Systematic screening for active tuberculosis. World Health Organization (2013).

5. Migliori, G. B. et al. Reducing tuberculosis transmission: a consensus document from the World Health Organization Regional Office for Europe. Eur. Respir. J. 53, 1–18 (2019).

6. Gill, C. M., Dolan, L., Piggott, L. M. & McLaughlin, A. M. New developments in tuberculosis diagnosis and treatment. Breathe 18, 1–15 (2022).

7. Chung, K. F. & Pavord, I. D. Prevalence, pathogenesis, and causes of chronic cough. Lancet 371, 1364–1374 (2008).

8. Simonsson, B. G., Jacobs, F. M. & Nadel, J. A. Role of Autonomic Nervous System and the Cough Reflex in the Increased Responsiveness of Airways in Patients with Obstructive Airway Disease. J. Clin. Invest. 46, 1812–1818 (1967).

9. Turner, R. D. & Bothamley, G. H. Cough and the Transmission of Tuberculosis. J. Infect. Dis. 211, 1367–1372 (2015).

10. Swarnkar, V. et al. Automatic Identification of Wet and Dry Cough in Pediatric Patients with Respiratory Diseases. Ann. Biomed. Eng. 41, 1016–1028 (2013).

11. Kaplan, A. G. Chronic Cough in Adults: Make the Diagnosis and Make a Difference. Pulm. Ther. 5, 11–21 (2019).

12. Proaño, A. et al. Protocol for studying cough frequency in people with pulmonary tuberculosis. BMJ Open 6, 1–9 (2016).

13. Botha, G. H. R. et al. Detection of tuberculosis by automatic cough sound analysis. Physiol. Meas. 39, 045005 (2018).

14. Pahar, M. et al. Automatic cough classification for tuberculosis screening in a real-world environment. Physiol. Meas. 42, 105014 (2021).

15. Armstrong, S. The computer will assess you now. BMJ 355, 1–2 (2016).

16. The Lancet. Artificial intelligence in health care: within touching distance. Lancet 390, 2739 (2017).

17. Topalovic, M. et al. Artificial intelligence outperforms pulmonologists in the interpretation of pulmonary function tests. Eur. Respir. J. 53, 1–11 (2019).

18. Ijaz, A. et al. Towards using cough for respiratory disease diagnosis by leveraging Artificial Intelligence: A survey. Informatics Med. Unlocked 29, 1–28 (2022).

19. Kik, S. V., Denkinger, C. M., Casenghi, M., Vadnais, C. & Pai, M. Tuberculosis diagnostics: which target product profiles should be prioritised? Eur. Respir. J. 44, 537–540 (2014).

20. Larson, S. et al. Validation of an Automated Cough Detection Algorithm for Tracking Recovery of Pulmonary Tuberculosis Patients. PLoS One 7, 1–10 (2012).

21. Pourhoseingholi, M. A., Vahedi, M. & Rahimzadeh, M. Sample size calculation in medical studies. Gastroenterol. Hepatol. from Bed to Bench 6, 14–17 (2013).

22. National TB elimination programme Central TB Division. National TB Prevalence Survey in India 2019 - 2021. Ministry of Health and Family Welfare https://tbcindia.gov.in/showfile.php?lid=3659 (2021).

23. Jones, P. W., Quirk, F. H. & Baveystock, C. M. The St George’s Respiratory Questionnaire. Respir. Med. 85, 25–31 (1991).

24. Barry, S. J., Dane, A. D., Morice, A. H. & Walmsley, A. D. The automatic recognition and counting of cough. Cough 2, 8 (2006).

25. Rudraraju, G. et al. Cough sound analysis and objective correlation with spirometry and clinical diagnosis. Informatics Med. Unlocked 19, 1–11 (2020).

26. Ribeiro, M., Singh, S. & Guestrin, C. “Why Should I Trust You?”: Explaining the Predictions of Any Classifier. in Proceedings of the 2016 Conference of the North American Chapter of the Association for Computational Linguistics: Demonstrations 1135–1144 (Association for Computational Linguistics, 2016). doi:10.18653/v1/N16-3020.

27. Clopper, C. J. & Pearson, E. S. The Use of Confidence or Fiducial Limits Illustrated in the Case of the Binomial. Biometrika 26, 404–413 (1934).

28. Belkacem, A. N., Ouhbi, S., Lakas, A., Benkhelifa, E. & Chen, C. End-to-End AI-Based Point-of-Care Diagnosis System for Classifying Respiratory Illnesses and Early Detection of COVID-19: A Theoretical Framework. Front. Med. 8, 1–13 (2021).

29. Abeyratne, U. R., Swarnkar, V., Setyati, A. & Triasih, R. Cough Sound Analysis Can Rapidly Diagnose Childhood Pneumonia. Ann. Biomed. Eng. 41, 2448–2462 (2013).

30. Xu, X. et al. Listen2Cough: Leveraging End-to-End Deep Learning Cough Detection Model to Enhance Lung Health Assessment Using Passively Sensed Audio. Proc. ACM Interactive, Mobile, Wearable Ubiquitous Technol. 5, 1–22 (2021).

31. Klco, P., Kollarik, M. & Tatar, M. Novel computer algorithm for cough monitoring based on octonions. Respir. Physiol. Neurobiol. 257, 36–41 (2018).

32. Laguarta, J., Hueto, F. & Subirana, B. COVID-19 Artificial Intelligence Diagnosis Using only Cough Recordings. IEEE Open J. Eng. Med. Biol. 1, 275–281 (2020).

33. Pahar, M., Klopper, M., Warren, R. & Niesler, T. COVID-19 cough classification using machine learning and global smartphone recordings. Comput. Biol. Med. 135, 1–10 (2021).

